# The Impact of Multi-Cancer Early Detection Tests on Cancer Mortality: A 10-Year Microsimulation Model

**DOI:** 10.64898/2026.05.05.26351205

**Authors:** Jade Xiao, Andrew K. ElHabr, Christopher Tyson, Xiting Cao, A. Mark Fendrick, A. Burak Ozbay, Paul Limburg, Tomasz M. Beer, Ashish A. Deshmukh, Jagpreet Chhatwal

**Author notes:** **Corresponding Author**, Jagpreet Chhatwal, PhD, MGH Institute for Technology Assessment, 125 Nashua St, Suite 660, Boston, MA 02114.

## Abstract

**Purpose:** Early detection of cancer can improve survival following diagnosis. However, routine screening is limited to a few cancer types. Multi-cancer early detection (MCED) tests could substantially expand cancer screening by simultaneously detecting multiple cancer types. This modeling study evaluates the potential impact of an MCED test on cancer outcomes in the US general population.

**Methods:** We developed a microsimulation model of 14 solid tumor cancer types which account for nearly 80% of cancer incidence and mortality. The model was calibrated to reproduce annual incidence rates reported in the Surveillance, Epidemiology, and End Results database. Cancer diagnosis could arise from standard-of-care (SoC) procedures or annual MCED testing. MCED sensitivities were derived from a case-control clinical validation study. We simulated the 10-year life course of 5 million US adults aged 50-84 years. The primary outcome was cancer mortality reduction due to MCED testing.

**Results:** In the best case with perfect uptake and adherence, MCED testing added to the SoC led to a 23% decrease in 10-year cancer mortality relative to the SoC alone, translating to 668,600 cancer deaths averted over 10 years. The largest mortality reductions, in absolute terms, were observed for lung (160; 802 versus 962 per 100,000), colorectal (118; 168 versus 284), and pancreatic (50; 238 versus 288) cancer. The largest relative reductions were in cervical (52%), colorectal (41%), and breast (34%) cancer. The population-level life-year gain was 7,158 years per 100,000.

**Conclusion:** MCED testing has the potential to substantially reduce cancer-related deaths, improve outcomes across multiple cancer types.

## INTRODUCTION

Over the last two decades, cancer mortality in the United States (US) has seen drastic declines––34% relative reduction from 1991 to 2023, translating to a reduction of 4.8 million cancer deaths––driven by decreased rates of smoking and increased uptake of screening alongside therapeutic advances.^1^ Despite this progress, cancer remains a leading cause of death. Projections for 2026 estimate that there will be more than 2.1 million new cancer diagnoses and 626,140 cancer deaths in the US.^1^ Early detection of cancer has been linked to improved survival by averting progression to metastasis,^2^ yet most cancer types do not have currently recommended screening strategies. Cancer types excluding the four with routine screening recommended by the United States Preventive Services Taskforce (USPSTF) (i.e., breast, cervical, colorectal, and lung),^3^ collectively account for 70% of new cancer diagnoses.^4^ Given that around half of cancer cases in the US are detected at an advanced stage,^5^ there is great potential for further mortality reductions to be realized through early detection.

Liquid biopsy-based multi-cancer early detection (MCED) tests continue to demonstrate their potential to revolutionalize early cancer detection. These tests are capable of detecting cancer signals from multiple cancer types simultaneously. Several MCED tests have been evaluated in clinical trials. The **D**etecting cancers **E**arlier **T**hrough **E**lective mutation-based blood **C**ollection and **T**esting (DETECT-A) study was the the first large, prospective, interventional clinical trial of an MCED test followed by PET-CT triage. It found that all eight patients with stage I-II MCED-detected cancer who underwent surgery remained cancer-free after the median follow-up of 4.4 years.^6,7^ Subsequently, a large, multi-center, prospective, case-control study, **A**scertaining **S**erial **C**ancer patients to **E**nable **N**ew **D**iagnostic 2 (ASCEND-2), demonstrated a test sensitivity of 50.9% and specificity of 98.5% across 21 cancer types.^8^ Most recently, the clinical performance of a commercially available MCED test was evaluated in a case-control clinical validation study using peripheral blood samples selected with consideration to the characteristics of the intended use population. The overall sensitivity was 57.8% (64.1% excluding breast and prostate cancer) and specificity was 97.4%.^9^

Given that the real-world impact of MCED tests will not be known for many years, simulation modeling can provide interim projections of cancer outcomes following their introduction. In particular, overdiagnosis—the detection of cancers that would not otherwise have resulted in symptoms or harm––has been a long-standing concern surrounding cancer screening as it causes unnecessary treatments and patient distress. This concern understandably extends to MCED tests. Simulation modeling can also estimate overdiagnosis rates caused by MCED testing to address these concerns. The objective of this study was to evaluate the potential benefits of incorporating an MCED test into the standard of care (SoC) for the US general population. To that end, we developed a microsimulation model of 14 cancer types that collectively account for nearly 80% of all incident cancers.^10^ The primary outcomes were cancer mortality reduction and survival gains due to stage shift––the downward shift in cancer stage at the point of diagnosis––relative to the SoC.

## MATERIALS & METHODS

We extended a previously-validated discrete-event microsimulation model, the **Si**mulation Model for **MCED** (SiMCED),^11^ of 14 solid tumor cancer types: breast, cervical, colorectal, endometrial, esophageal, gastric, head and neck, kidney, liver, lung, ovarian, pancreatic, prostate, and urinary bladder.

### Simulated Cohort

The simulated cohort consisted of 5 million adults aged 50-84 years without a cancer diagnosis, representing approximately 95-100 milllion US adults in 2015. The composition of sex, race, and age was consistent with that of the US population in 2015.^12^

### Natural History

A simulated individual can develop only one out of the 14 included cancer types in their lifetime. For each cancer type, oncogenesis occurs at a rate specific to the individual’s sex, race, and age. The cancer type with the earliest time of oncogenesis before the time of death is the cancer type that the individual develops. In the absence of a diagnosis, cancer progresses according to cancer type- and stage-specific dwell times synthesized from published literature and expert surveys (**Table S3**).^13,14^

### Cancer Diagnosis

Diagnosis under the SoC encompasses existing routine screening procedures, incidental detection, and symptomatic presentation. Diagnosis was assumed to occur immediately upon advancement to stage IV cancer due to the relatively short pre-symptomatic phase. In all other stages, SoC diagnosis occurs at a rate specific to the cancer type and stage, as well as the individual’s sex, race, and age. MCED testing was modeled as a supplemental screening approach with cancer type- and stage-specific sensitivities derived from a case-control clinical validation study.^9^ The base case represents the best-case scenario, where the MCED test is administered annually at the beginning of each calendar year to individuals aged 50-84 years, with the assumption of 100% uptake (i.e., the proportion of the cohort who will take the MCED test at all) and 100% adherence (i.e., the probability of an individual accepting the MCED test each time it is offered). It is unclear what impact, if any, MCED testing will have on real-world SoC screening uptake and adherence. Nevertheless, the MCED test is intended to supplement––not replace––existing screening practices. For these reasons, we hypothesized that the introduction of MCED testing would have no effect on SoC screening. Scenarios with decreased or increased rates of SoC diagnosis were not explored. Follow-up evaluation after a positive MCED test result was assumed to be 100%.

### Model Analysis

Using the same population of individuals, the model was run twice, once without MCED (“SoC”) and once with MCED (“SoC + MCED”). Thus, the same individual across both instances formed a pair of digital twins, such that differences in outcomes can be directly attributed to MCED testing.

### Cancer Survival

Mortality arises from two sources: (1) non-cancer background mortality and (2) cancer-specific mortality following diagnosis. Background mortality was modeled according to US life tables stratified by sex, age, and race.^15^ Individuals could die from non-cancer causes at any time, regardless of cancer status. Cancer-specific mortality was applied only after diagnosis, i.e., individuals with undiagnosed cancer were not permitted to die of cancer. After diagnosis, individuals followed cancer type- and stage-specific survival curves to determine the time and cause of death.^10^ By design, MCED testing can only advance the time of diagnosis. However, naïve application of survival curves from the point of MCED detection could introduce artificial harm, i.e., earlier death despite earlier diagnosis due to the additional mortality risk from cancer post diagnosis. To preserve clinical face validity, we imposed the following rules to model cancer survival after MCED detection.

For each individual in “SoC + MCED”, their time and cause of death was evaluated with reference to the time and cause of death of their digital twin in “SoC” (**Figure 1**). If the cause of death in “SoC” was non-cancer, then the individual is assigned the same time of non-cancer death in “SoC + MCED”. Thus, earlier cancer detection was not permitted to alter non-cancer mortality. Conversely, if the cause of death in “SoC” was cancer, then the individual remains alive until at least the time of cancer death in “SoC”, at which point they begin following survival curves corresponding to their stage at MCED detection. In other words, the individual is guaranteed to survive the initial portion of the survival curve given by the number of years between MCED detection in “SoC + MCED” and cancer death in “SoC”. Operationally, this approach delays the risk of the death in “SoC + MCED” until the time of death in “SoC” to ensure that modeled mortality benefits from earlier diagnosis are not influenced by lead-time artifacts.

**Figure 1.**
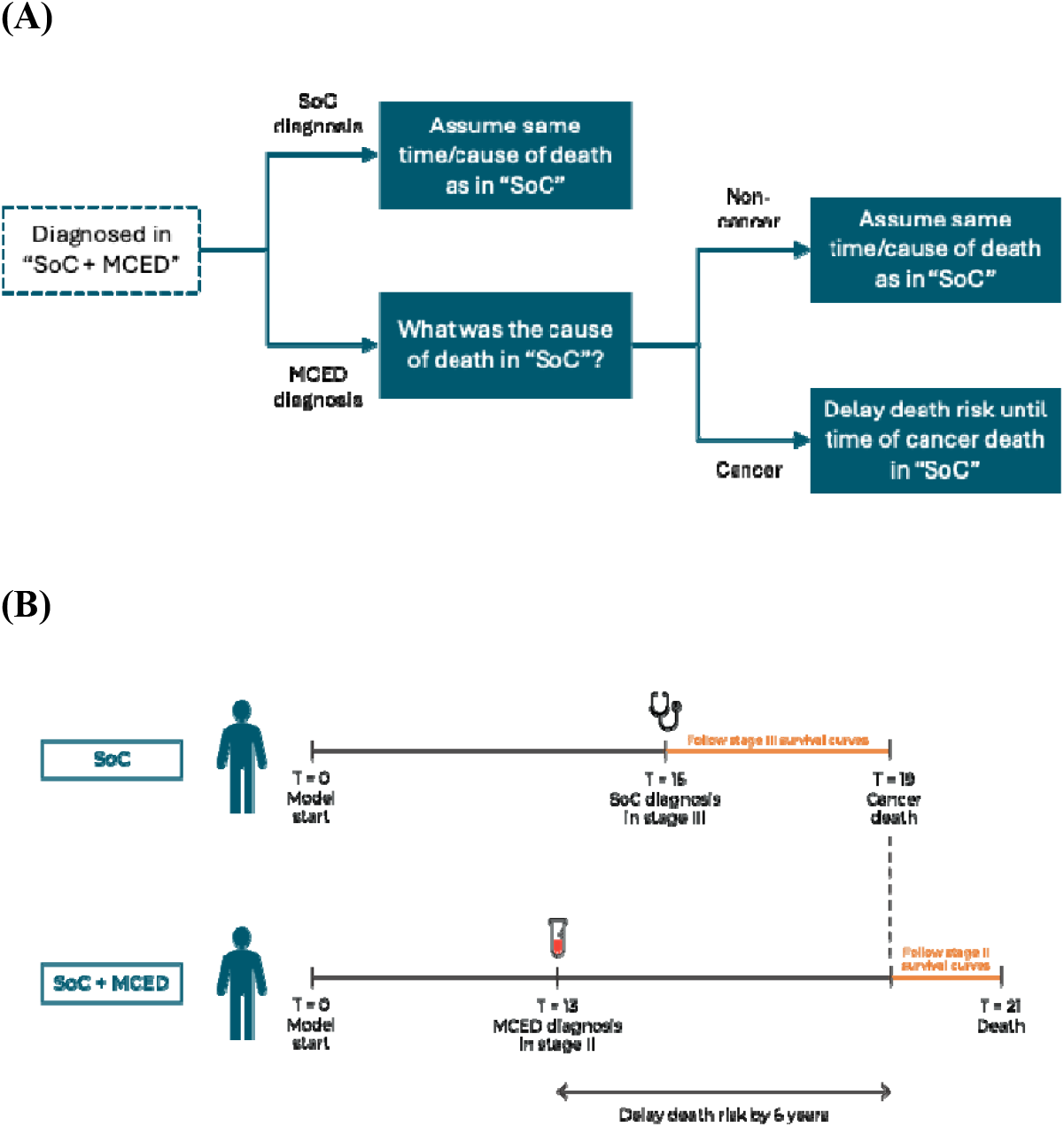
**(A)** Post diagnosis mortality decision tree. **(B)** Example application. Abbreviations: MCED, multi-cancer early detection, SoC, standard of care.

### Model Outcomes

The main result was stage shift, cancer mortality reduction, and life-years gained (LYG) due to the supplemental use of an MCED test over a time horizon of 10 years. We report all incidences as rates per an initial closed cohort of 100,000. For individuals with SoC diagnosis within the 10-year horizon, LYG was estimated at the individual- and population-levels, with the latter being defined as the total LYG in an initial closed cohort of 100,000. Due to the uncertain validity of extrapolated survival curves in the long term, the LYG from MCED detection is voided if the survival in “SoC” is greater than 10 years. In addition, LYG is capped at 10 years.

### Scenario Analyses

To evaluate the robustness and sensitivity of our model assumptions and conclusions, we simulated various scenarios. First, we replaced annual testing with biennial and triennial testing. Second, we investigated imperfect uptake and adherence levels of 90%, 70%, and 50%. The lower bound of 50% is below the real-world adherence to non-invasive laboratory tests for cancer detection, such as those for colorectal cancer.^16–18^ Third, we modeled reduced MCED sensitivities by applying a discount factor of 80% to account for potentially lower effectiveness in a real-world setting. Fourth, we varied dwell times by ±25% to model different speeds of progression. Unobserved incidence was not included separately in the scenario analysis. Fifth, we simulated a scenario with one-time MCED testing in Year 1 to isolate the effect of a single MCED test over a 10-year period. Lastly, we tested different approaches to modeling survival after diagnosis. Instead of delaying the risk of death in “SoC + MCED” until the time of death in “SoC”, we implemented (1) delaying the risk of death until only the time of diagnosis in “SoC”, and (2) no delaying the risk of death. These alternative approaches, though relying on fewer assumptions, have reduced clinical face validity as they allow for the possibility of earlier death in “SoC + MCED”.

## RESULTS

Over the 10-year horizon, supplemental testing with an annual MCED test with perfect uptake and adherence resulted in a 14% increase (3,502 versus 3,068 cases per 100,000) in stage I diagnoses, 27% increase (2,642 versus 2,079) in stage II diagnoses, and 29% increase (1,826 versus 1,414) in stage III diagnoses, relative to the SoC alone; in contrast, stage IV diagnoses decreased by 51% (1,032 versus 2,108) (**Figures 2A** and **S4**). The increase in stage III diagnoses is due to the relatively low MCED sensitivities in stages I-II and relatively high sensitivities in stage III, causing downstaging cases from stage IV to III to outnumber those from stage III to I-II. The cumulative number of diagnoses was 8,669 under the SoC, and 9,002 when supplemented by MCED testing, equating to a modest increase of 3.8% (333 per 100,000). Of these 333 additional diagnoses, 218 were made in individuals who died from non-cancer-related causes under the SoC after their counterfactual time of MCED detection, and 115 were in individuals who were eventually diagnosed under the SoC after the first 10 years. **Figure 2B** depicts the flow of individuals from their stage-at-diagnosis under the SoC to their stage-at-diagnosis when SoC is supplemented with MCED testing.

**Figure 2.**
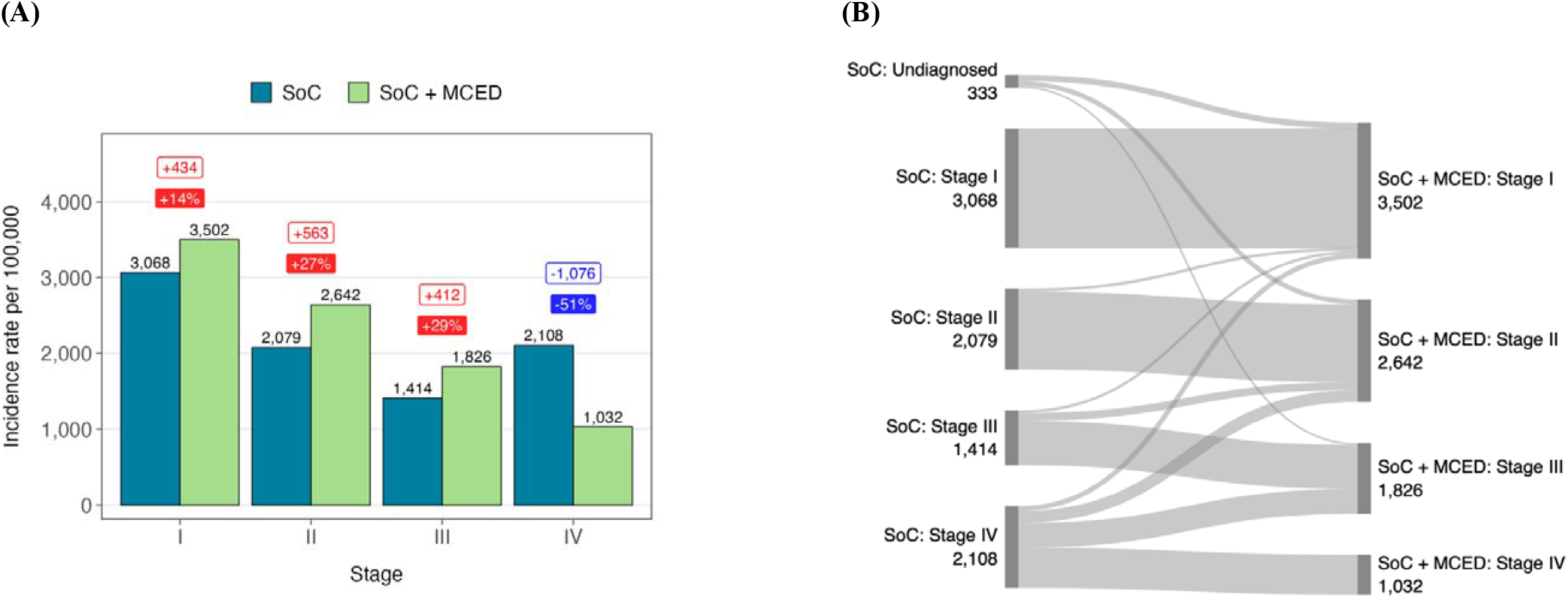
10-year **(A)** stage shift and **(B)** individual-level downstaging flows. Numbers are counts of individuals per 100,000. Note that “SoC: Undiagnosed” does not depict all undiagnosed cases, only those who are undiagnosed in the “SoC” scenario but become diagnosed in the “SoC + MCED” scenario within the 10-year time horizon. Abbreviations: MCED, multi-cancer early detection; SoC, standard of care.

Cancer-related deaths over this time period decreased 23% from 2,537 to 1,965 per 100,000 (**Table 1**). The cancer types with the largest absolute reductions in mortality per 100,000 were, in order, lung (160; 802 versus 962), colorectal (118; 168 versus 284), and pancreatic (50; 238 versus 288). The cancer types with the largest relative reductions were cervical (52%), colorectal (41%), and breast (34%). Mortality reduction was higher for cancer types with recommended screening (24%) than for those without recommended screening (21%).

**Table 1.** 10-year cancer mortality by cancer type.

| **Cancer type** | **SoC** | **SoC +**<br>**MCED** | **Absolute**<br>**change** | **Relative**<br>**change** |
| --- | --- | --- | --- | --- |
| Breast | 120 | 79 | -41 | -34% |
| Cervical | 18 | 9 | -9 | -52% |
| Colorectal | 284 | 168 | -116 | -41% |
| Endometrial | 60 | 47 | -13 | -22% |
| Esophageal | 83 | 69 | -14 | -17% |
| Gastric | 105 | 73 | -32 | -30% |
| Head and Neck | 114 | 79 | -35 | -31% |
| Kidney | 89 | 63 | -26 | -29% |
| Liver | 177 | 141 | -36 | -20% |
| Lung | 962 | 802 | -160 | -17% |
| Ovarian | 64 | 56 | -8 | -12% |
| Pancreatic | 288 | 238 | -51 | -18% |
| Prostate | 79 | 77 | -2 | -2% |
| Urinary Bladder | 94 | 64 | -30 | -32% |
| **Total** | **2,537** | **1,965** | **-572** | **-23%** |
| **Total for cancer types**<br>**with recommended**<br>**screening** | **1,384** | **1,058** | **-326** | **-24%** |
| **Total for cancer types**<br>**without recommended**<br>**screening** | **1,153** | **907** | **-246** | **-21%** |

Abbreviations: MCED, multi-cancer early detection; SoC, standard of care.

Figure 3 breaks down LYG by downstaging flow. Overall, 16.5% of cancer cases were downstaged (**Figure 3A**), i.e., the stage at diagnosis in “SoC + MCED” was strictly lower than the stage at diagnosis in “SoC”, and a further 9.6% were diagnosed earlier in the same stage (2.0% in stage I, 3.3% in stage II, and 4.4% in stage III). In general, the more dramatic the downstaging, the greater the LYG (**Figure 3B**). When downstaged from stage IV to III, II, and I, the mean LYG was 3.7, 5.1, and 6.0 years, respectively. When downstaged from stage III to II and I, the mean LYG was 2.7 and 4.1 years, respectively. For additional detail, **Table S4** describes the distribution of LYG in terms of the proportion of cases exceeding certain LYG thresholds. Population-level LYG reflects both the frequency of the downstaging flow and the associated LYG (**Figure 3C**). The most common downstaging flow of stage IV to III translates to a population-level LYG of 2,381 years per 100,000, followed by 1,559 for stage IV to II, and 722 for stage IV to I. Among patients diagnosed earlier in the same stage, mean LYGs were below 1 at the individual level, but the aggregate LYG in a population of 100,000 was 197 for stage I, 413 for stage II, and 818 for stage III. The total population-level LYG was 7,158 years per 100,000.

**Figure 3.**
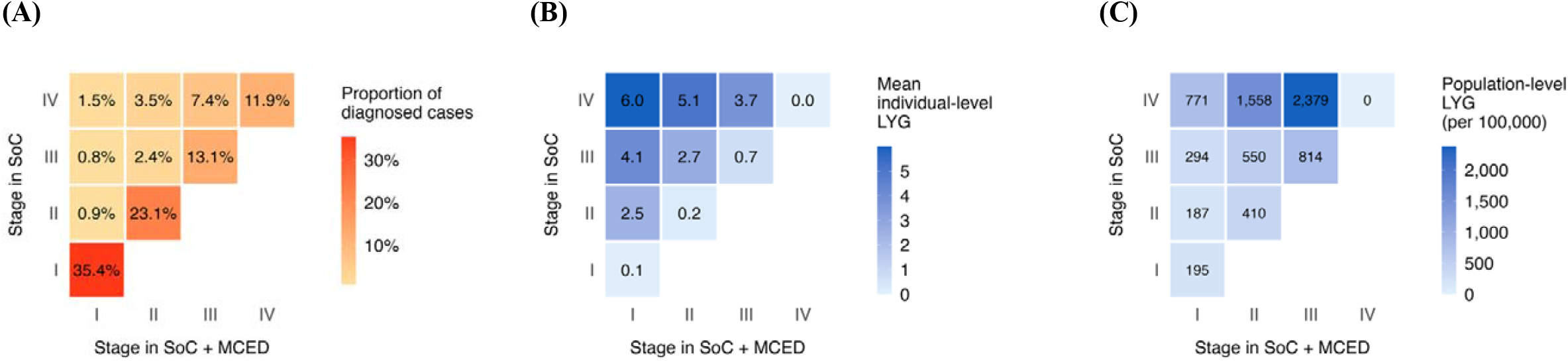
**(A)** Proportion of diagnosed cases, **(B)** individual-level LYG, and **(C)** population-level LYG by downstaging flow. Abbreviations: LYG, life-years gained.

**Table 2** summarizes the findings from the scenario analyses, where MCED testing interval, MCED uptake and adherence, MCED sensitivities, and cancer dwell times were varied within clinically plausible ranges. Overall, testing interval had the greatest impact on mortality reduction. Longer testing intervals were associated with smaller mortality reductions, which dropped steeply from 51% in the base case with annual testing (10 total tests over 10 years) to 33% with biennial testing (5 total tests) and 26% with triennial testing (4 total tests). Reducing MCED uptake and adherence to 90% produced a modest change in outcomes, achieving 46% and 47% mortality reduction, respectively. Reducing uptake and adherence to 70% produced mortality reductions of 36% and 38%, respectively. Reducing uptake and adherence to 50% produced mortality reductions of 25% and 28%, respectively. Applying a discount factor of 80% to MCED sensitivities resulted in a mortality reduction of 18%. In the scenario with one-time MCED testing in Year 1, mortality reduction was only 4%.

**Table 2.** 10-year cancer mortality by scenario.

| Scenario name | SoC | SoC +<br>MCED | Absolute<br>change | Relative<br>change |
| --- | --- | --- | --- | --- |
| **Base case** |  |  |  |  |
|  | 2,537 | 1,965 | -572 | -23% |
| **MCED testing interval** |  |  |  |  |
| Biennial | 2,537 | 2,164 | -373 | -15% |
| Triennial | 2,537 | 2,261 | -275 | -11% |
| **MCED uptake** |  |  |  |  |
| 90% | 2,537 | 2,022 | -515 | -20% |
| 70% | 2,537 | 2,134 | -402 | -16% |
| 50% | 2,537 | 2,251 | -286 | -11% |
| **MCED adherence** |  |  |  |  |
| 90% | 2,537 | 2,013 | -524 | -21% |
| 70% | 2,537 | 2,115 | -421 | -17% |
| 50% | 2,537 | 2,229 | -308 | -12% |
| **MCED sensitivities discounting** |  |  |  |  |
| 80% | 2,537 | 2,088 | -449 | -18% |
| **Dwell times** |  |  |  |  |
| Slower | 2,555 | 1,945 | -609 | -24% |
| Faster | 2,714 | 2,143 | -571 | -21% |
| **One-time MCED testing** |  |  |  |  |
|  | 2,537 | 2,438 | -99 | -4% |
| **Survival approach** |  |  |  |  |
| Delay death risk until time of “SoC” diagnosis | 2,537 | 2,186 | -351 | -14% |
| No delay of death risk | 2,537 | 2,338 | -198 | -8% |

Abbreviations: MCED, multi-cancer early detection; SoC, standard of care.

As expected, the scenario with slower dwell times had better mortality reduction (24%) compared to the base case due to greater chances of MCED detection when a cancer progresses more slowly. Conversely, the scenario with faster dwell times had worse mortality reduction (21%) due to reduced opportunities for MCED detection with a faster progressing cancer. Mortality reduction was sensitive to the survival approach. When the risk of death was delayed only until the “SoC” time of diagnosis, mortality reduction fell to 14%. When no delay was enforced, mortality reduction was 8%. It should be noted that, while these alternative approaches rely on fewer assumptions and still lead to mortality reduction at an aggregate level, they have limited face validity at the individual level since an earlier diagnosis can lead to earlier death, reducing the utility of early cancer detection.

## DISCUSSION

In this microsimulation analysis of 14 major solid tumor cancer types, we predicted that annual MCED testing with perfect uptake and adherence, when added to current standard of care screening, could meaningfully shift the stage at diagnosis and reduce cancer mortality over a 10-year period. In the base case, annual testing resulted in a 23% reduction in cancer-related deaths. Projected onto the US population aged 50-84 years,^19^ this translates to 668,600 cancer deaths averted over 10 years. Mortality reductions were observed across both cancers with and without established screening programs, with particularly large absolute reductions for lung, colorectal, and pancreatic cancer.

Our study adds to the growing body of literature on simulation modeling for MCED testing.^20^ Hubbell et al. developed a model of 25 cancer types in the US population aged 50-79 years. Annual testing reduced cancer-related deaths by 26%, equivalent to 104 per 100,000.^21^ Kansal et al. evaluated annual MCED testing with 90% adherence in a cohort of US individuals agd 50-79 years.^22^ The mean LYG was 4.85 years per downstaged patient, and 0.18 years when averaged over the entire cohort. Gogebakan et al. adapted the 12-cancer model by Lange et al. for the average-risk population in England, with a median age of 66 years. Assuming 100% adherence to 3 rounds of annual testing, the 5-year cancer mortality reduction was 6-9%.^23,24^ While our findings complement these studies, the key strength of our model is its ability to estimate overdiagnosis as a direct consequence of MCED testing arising naturally from the MCED sensitivities. This is in contrast to Kansal et al., who treated the overdiagnosis rate as an external model parameter.^22^ Our results indicate that the increase in total diagnoses was only 3.8% with the addition of MCED testing, suggesting that overdiagnosis may not be an issue with this technology. This is a critical finding as it mitigates concerns that MCED testing could lead to a surge in unnecessary cancer diagnoses and treatment. Additionally, this analysis includes a comprehensive scenario analysis to examine the effect of different testing intervals, uptake rates, and adherence levels.

The key finding of our study is the potential 23% reduction in 10-year cancer mortality and population-level LYG of 7,158 years per 100,000 due to annual MCED testing with perfect uptake and adherence. Our breakdown of mortality reduction by cancer type further emphasizes the clinical utility of MCED testing as the cancer types that had the largest absolute reductions (i.e., lung, colorectal, and pancreatic) are among the most aggressive cancer types with the poorest survival rates. Interestingly, mortality reduction was greater for cancer types with recommended screening than for those without. This suggests that MCED testing could be effective for boosting mortality reduction when used to supplement routine screening, while being the main driver of this outcome for cancer types without screening tests.

In our scenario analysis, testing frequency significantly impacted mortality reduction. Biennial and triennial testing achieved mortality reductions of only 15% and 11%, respectively. One-time MCED testing at the beginning of the 10-year period produced a modest mortality reduction of 4%. These results indicate that frequent testing is an important factor determining the real-world effectiveness of MCED tests. Our study also explored the impact of uptake and adherence. Even under scenarios where uptake and adherence were reduced to 50%, MCED testing still resulted in 11-12% mortality reduction. While this is a positive outcome, it underscores the importance of public health initiatives––such as public awareness campaigns, education on the benefits of early cancer detection, and efforts to reduce disparities in access to screening––to promote widespread adoption and consistent use of MCED tests.

We recognize several limitations of this research. First, the model does not account for the potentially poorer prognosis of MCED-detected cancers. There is emerging evidence that tumor DNA shedding, an important determinant of cancer signal detection by MCED tests, is also a signifier of worse prognosis.^25,26^ A second limitation is also related to model structure. Routine screening for the four USPTSF-recommended cancers is not modeled explicitly but captured implicitly via diagnosis rates. These rates were assumed to remain the same after the introduction of an MCED test. It is possible that the availability of MCED testing may cause individuals to forgo routine screening or to reduce their screening frequency. Conversely, MCED testing could also increase screening rates by promoting awareness of cancer screening. Thus far, neither effect has been reliably established in published studies. Furthermore, the MCED test was intended to supplement––not replace––SoC screening; therefore, scenarios with decreased or increased rates of SoC diagnosis were not explored. Third, our model does not allow individuals to develop more than one cancer type in their lifetime, although the MCED test is capable of detecting second primary or recurrent cases. Therefore, the true benefit of MCED testing may be underestimated in this analysis. Another source of uncertainty is the performance of the MCED test in the real world, which may be lower than what was estimated in the case-control clinical validation study.^9^ We addressed this by simulating a scenario with sensitivity discounting, finding the stage shift benefit to be reduced but not insignificant.

## CONCLUSION

Our study shows that annual MCED testing with high rates of uptake, adherence, and follow-up evaluation after a positive result has the potential to substantially reduce cancer mortality, particularly for cancer types that lack routine screening programs. While further research is needed to validate these findings in real-world settings, our results suggest that MCED testing could transform cancer diagnosis and improve patient outcomes across a broad range of cancer types.

## Data Availability

All data produced are available on request.

## Conflict of Interest Statement

Chhatwal has ownership in Value Analytics Labs. Tyson, Cao, Ozbay, Limburg, and Beer are employees of Abbott Laboratories (formerly Exact Sciences) and hold stock in the company. Fendrick directs the University of Michigan Center for Value-Based Insurance Design. He reports providing consulting services to AbbVie, CareFirst Blue Cross Blue Shield, Centivo, Clover Health, Community Oncology Association, Covered California, Elektra Health, EmblemHealth, Employee Benefit Research Institute, Exact Sciences, GRAIL, Health[at]Scale Technologies, HealthCorum, MedZed Inc., Merck and Company, Mother Goose Health, Phathom Pharmaceuticals, Proton Intelligence, Inc., Sempre Health, Silver Fern Healthcare, US Department of Defense, Virginia Center for Health Innovation, Wellth, Yale-New Haven Health System; receiving research support from the Agency for Healthcare Research and Quality, West Health Policy Center, Arnold Ventures, National Pharmaceutical Council, Patient-Centered Outcomes Research Institute, Pharmaceutical Research and Manufacturers of America, the Robert Wood Johnson Foundation, the state of Michigan, and the Centers for Medicare and Medicaid Services; serving as coeditor for the *American Journal of Managed Care*; and maintaining a partnership at VBID Health. Deshmukh is a consultant for Merck Inc. and Value Analytics Labs. Deshmukh received consulting fees from Value Analytics Labs. No additional conflicts of interest were reported by the rest of the authors.

## Data Availability Statement

All data generated by the simulation model are made available in the manuscript. Model code is unable to shared publicly due to proprietary restrictions.

## Funding

This study was funded by Exact Sciences Corporation, Madison, WI.

## Notes

### Author Declarations

The study used ONLY openly available aggregated human data from published sources.

## REFERENCES

1. Siegel RL, Kratzer TB, Wagle NS, Sung H, Jemal A. Cancer statistics, 2026. CA Cancer J Clin. 2026;76(1):e70043. doi:10.3322/caac.70043

2. Yu M, Tyson C, Limburg PJ, Beer TM. A flexible quantitative framework to assess the potential contribution of early cancer detection to improved cancer survival. J Clin Oncol. 2023;41(16_suppl):e22508–e22508. doi:10.1200/JCO.2023.41.16_suppl.e22508

3. Centers for Disease Control and Prevention (CDC). Cancer Screening Tests. October 17, 2023. Accessed September 25, 2023. https://www.cdc.gov/cancer/prevention/screening.html

4. Siegel RL, Giaquinto AN, Jemal A. Cancer statistics, 2024. CA Cancer J Clin. 2024;74(1):12–49. doi:10.3322/caac.21820

5. Crosby D, Bhatia S, Brindle KM, et al. Early detection of cancer. Science. 2022;375(6586):eaay9040. doi:10.1126/science.aay9040

6. Buchanan AH, Lennon AM, Choudhry OA, et al. Multiyear Clinical Outcomes of Cancers Diagnosed Following Detection by a Blood-Based Multicancer Early Detection Test. Cancer Prev Res (Phila Pa). 2024;17(8):349–353. doi:10.1158/1940-6207.CAPR-24-0107

7. Lennon AM, Buchanan AH, Rego SP, et al. Outcomes Following a False-Positive Multi-Cancer Early Detection Test: Results from DETECT-A, the First Large, Prospective, Interventional MCED Study. Cancer Prev Res (Phila Pa). 2024;17(8):355–359. doi:10.1158/1940-6207.CAPR-23-0451

8. Gainullin V, Bae J, Guthrie VB, et al. Abstract A056: Performance of multi-biomarker class reflex testing in a prospectively-collected cohort. Clin Cancer Res. 2024;30(21_Supplement):A056. doi:10.1158/1557-3265.LIQBIOP24-A056

9. Gainullin VG, Gray M, Kumar M, et al. Performance of an Optimized Methylation-Protein Multi-Cancer Early Detection (MCED) Test Classifier. medRxiv. Preprint posted online March 19, 2026:2026.03.03.26347329. doi:10.64898/2026.03.03.26347329

10. National Cancer Institute, DCCPS, Surveillance Research Program. Surveillance Research Program, National Cancer Institute SEER*Stat software. Published online November 2023. seer.cancer.gov/seerstat

11. Chhatwal J, Xiao J, ElHabr AK, et al. The impact of multicancer early detection tests on cancer stage shift: A 10-year microsimulation model. Cancer. 2025;131(22):e70075. doi:10.1002/cncr.70075

12. Centers for Disease Control and Prevention (CDC). Single-Race Population Estimates. https://wonder.cdc.gov/single-race-population.html

13. Broder MS, Ailawadhi S, Beltran H, et al. Estimates of stage-specific preclinical sojourn time across 21 cancer types. J Clin Oncol. 2021;39(15_suppl):e18584–e18584. doi:10.1200/JCO.2021.39.15_suppl.e18584

14. Shah N, Hathaway C, Tyson C, Cohain A, Li Y. NOVEL EMPIRICAL METHODS TO DERIVE STAGE-SPECIFIC DWELL TIME AND IMPLICATIONS FOR MULTI-CANCER EARLY DETECTION (MCED) MODELING.

15. Arias E, Xu J, Tejada-Vera B, Bastian B. U.S. State Life Tables, 2021. Natl Vital Stat Rep. 2024;73(7).

16. Greene M, Pew T, Dore M, et al. Re-screening adherence to multi-target stool DNA test for colorectal cancer: real-world study in a large national population. Int J Colorectal Dis. 2025;40(1):48. doi:10.1007/s00384-025-04837-6

17. Greene M, Pew T, Zapatier J, Rincón López JV, Limburg P, Duarte M. Adherence to Repeat Screening Completion for Colorectal Cancer Using the Multi-Target Stool DNA Test: Real-World Analysis of Patients from Federally Qualified Health Centers. J Prim Care Community Health. 2025;16:21501319251348099. doi:10.1177/21501319251348099

18. Le QA, Kiener T, Johnson HA, et al. Adherence to recommended blood-based screening tests for cancer and chronic diseases: A systematic literature review. Prev Med. 2025;191:108213. doi:10.1016/j.ypmed.2024.108213

19. United States Census Bureau. Annual Estimates of the Resident Population for Selected Age Groups by Sex for the United States: April 1, 2020 to July 1, 2024 (NC-EST2024-AGESEX). Published online January 2026. Accessed March 12, 2026. https://www2.census.gov/programs-surveys/popest/tables/2020-2024/national/asrh/nc-est2024-agesex.xlsx

20. Mandrik O, Whyte S, Kunst N, et al. Modeling the Impact of Multicancer Early Detection Tests: A Review of Natural History of Disease Models. Med Decis Mak Int J Soc Med Decis Mak. 2025;45(8):1013–1024. doi:10.1177/0272989X251351639

21. Hubbell E, Clarke CA, Aravanis AM, Berg CD. Modeled Reductions in Late-stage Cancer with a Multi-Cancer Early Detection Test. Cancer Epidemiol Biomark Prev Publ Am Assoc Cancer Res Cosponsored Am Soc Prev Oncol. 2021;30(3):460–468. doi:10.1158/1055-9965.EPI-20-1134

22. Kansal AR, Tafazzoli A, Shaul A, et al. Cost-effectiveness of a multicancer early detection test in the US. Am J Manag Care. 2024;30(12):e352–e358. doi:10.37765/ajmc.2024.89643

23. Gogebakan KC, Lange J, Owens L, et al. Clinical Significance of a Multicancer Screening Trial With Stage-Based End Points. JAMA Netw Open. 2025;8(10):e2536247. doi:10.1001/jamanetworkopen.2025.36247

24. Lange JM, Gogebakan KC, Gulati R, Etzioni R. Projecting the Impact of Multi-Cancer Early Detection on Late-Stage Incidence Using Multi-State Disease Modeling. Cancer Epidemiol Biomarkers Prev. 2024;33(6):830–837. doi:10.1158/1055-9965.EPI-23-1470

25. Chen X, Dong Z, Hubbell E, et al. Prognostic Significance of Blood-Based Multi-cancer Detection in Plasma Cell-Free DNA. Clin Cancer Res. 2021;27(15):4221–4229. doi:10.1158/1078-0432.CCR-21-0417

26. Bryce AH, Thiel DD, Seiden MV, et al. Performance of a Cell-Free DNA-Based Multi-cancer Detection Test in Individuals Presenting With Symptoms Suspicious for Cancers. JCO Precis Oncol. 2023;(7):e2200679. doi:10.1200/PO.22.00679

