## Supplementary Materials for "The Impact of Multi-Cancer Early Detection Tests on Cancer Mortality: A 10-Year Microsimulation Model"

**TECHNICAL SUPPLEMENT FOR SiMCED**

### S1 Overview

We developed **Si**mulation Model for **MCED** (SiMCED), a continuous-time, discrete-event microsimulation model of 14 solid tumor cancer types: breast, cervical, colorectal, endometrial, esophageal, gastric, head and neck, kidney, liver, lung, ovarian, pancreatic, prostate, and urinary bladder.^1^ These cancer types were selected based on the following reasons. First, they collectively account for nearly 80% of all incident cancers.^2^ Second, the model includes only cancer types that can be detected by the MCED test. Third, health states in SiMCED are based on the American Joint Committee on Cancer’s (AJCC’s) I–IV staging system for solid tumor cancer types; therefore, common blood-based cancers like leukemia, lymphoma, and myeloma were excluded due to incompatibility with the model structure. **Figure S1** is a high-level model schematic.

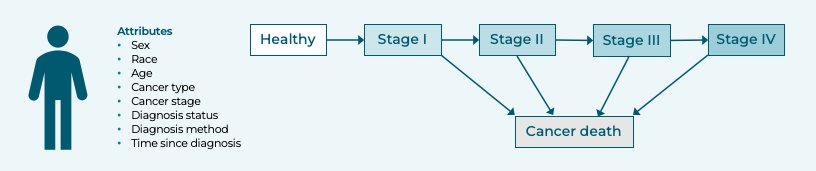

**Figure S1.** High-level model schematic of SiMCED.

A simulated individual can develop only one cancer type in their lifetime. Second primary and recurrent cancers are not modeled for the following reasons. First, they have markedly different pathogenesis and care processes that make them incompatible with the model structure. Second, the MCED test is not targeted towards individuals who have already had a cancer diagnosis and are undergoing surveillance for additional cancers.

The simulated cohort consists of U.S. adults aged 50–84 years (born 1931–1965) without a cancer diagnosis. Diagnosis of cancer can arise from SoC procedures or MCED testing. Diagnosis under the SoC encompasses existing routine screening procedures, incidental detection, and symptomatic presentation. MCED testing is modeled as a supplemental screening approach. Thus, the model is run twice, once without MCED (“SoC”) and once with MCED (“SoC + MCED”).

In the base case, the MCED test is administered annually at the beginning of each calendar year to individuals aged 50–84 years, with the assumption of 100% uptake (i.e., the proportion of the cohort who will take the MCED test at all) and 100% adherence (i.e., the probability of an individual accepting the MCED test each time it is offered). It is unclear what impact, if any, MCED testing will have on real-world SoC screening uptake and adherence. Nevertheless, the MCED test is intended to supplement––not replace––existing screening practices. For these reasons, we hypothesized that the introduction of MCED testing would have no effect on SoC screening.

### S2 Attributes

A simulated individual is initialized with the attributes in **Table S1**. Sex, race, and single year of age are jointly sampled from an empirical distribution representing the U.S. general population in 2015. This starting year was selected to allow for comparison against observed 5-year trends in cancer diagnosis.

**Table S1.** Individuals’ attributes.

| Attribute | Range |
| --- | --- |
| **Sex** | Female / Male |
| **Race** | Asian / Black / Hispanic / White / Others |
| **Age** | Single year of age between 50 and 84 |

Input data is stratified by attributes to the fullest extent possible (**Table S2**). Note that age stratification is at the 5-year age group level (50­–54, 55–59, 60–64, 65–69, 70–74, 75–79, 80–84, and 85+ years).

**Table S2.** Input data stratification.

| Input data | Sex | Age | Race | Cancer type | Cancer stage | Data source  (data year) | Calibration applied? |
| --- | --- | --- | --- | --- | --- | --- | --- |
| **Initial distribution of attributes** | ✓ | ✓ | ✓ | n/a | n/a | CDC WONDER^3^ (2015) | No |
| **Initial distribution of health states** | n/a | n/a | n/a | ✓ | ✓ | SEER (2010–2021)^2^ | Yes |
| **All-cause mortality** | ✓ | ✓ | ✓ | n/a | n/a | Arias et al.^4^ (2024) | No |
| **Rate of oncogenesis** | ✓ | ✓ | ✓ | ✓ | n/a | SEER (2010–2021)^2^ | Yes |
| **Rate of cancer progression** | x | x | x | ✓ | ✓ | Published literature^5,6^ | No |
| **Rate of SoC diagnosis** | ✓ | ✓ | ✓ | ✓ | ✓ | SEER (2010–2021)^2^ | Yes |
| **Cancer survival** | x | ✓ | x | ✓ | ✓ | SEER (2010–2021)^2^ | No |

Key: ✓ = full implementation; – = partial implementation; x = no implementation. Abbreviations: CDC, Centers for Disease Control and Prevention; MCED, multi-cancer early detection; n/a, not applicable; SEER, Surveillance, Epidemiology, and End Results; SoC, standard of care; WONDER, Wide-ranging ONline Data for Epidemiologic Research.

The following are justifications, where applicable, for not implementing full stratification.

##### Stratification of Cancer Survival

Raw survival curves are markedly different across age groups, but do not differ substantially across different sexes and races. Furthermore, SEER sample sizes become too small with stratification by sex, race, and age.

For a given (sex, race, cancer type, cancer stage), our criteria for using (sex, race, age group, cancer type, cancer stage)-stratified survival are:

1. The cohort size is at least 100 patients for all age groups;
2. There is at least 12 years of survival data for all age groups;
3. There is at least one cancer death for all age groups.

Under this criteria, full stratification is possible for only 0.5% of (sex, race, cancer type, cancer stage) combinations. For simplicity, we default to using only (age group, cancer type, cancer stage)-stratified survival.

### S3 Discrete Event Simulation

An individual’s life history is described by an “event list.” There are seven event types:

- Birth
- Oncogenesis
- Cancer progression
- SoC diagnosis
- MCED diagnosis
- Cancer death
- Non-cancer death

The following describes how the time to each event type is generated.

##### Non-Cancer Death

The age at non-cancer death is determined by sampling whether the individual survives each age after their age at initialization. This represents a “hard stop” for the individual’s life history, i.e., cancer can only cause the individual to die earlier than the time of non-cancer death. The maximum lifespan is 100 years.

##### Oncogenesis

The time to oncogenesis from the moment the individual attains their current 5-year age group is sampled from an exponential random variable with a rate given by the incidence rate for their (sex, race, age group), and resampled whenever the individual ages into the next age group. The cancer type with the earliest time of oncogenesis occurring before the time of non-cancer death is the cancer type that the individual develops.

After developing cancer, or if the individual initially has cancer, the next event is determined by whichever has the earliest time of occurrence among cancer progression, SoC diagnosis, MCED diagnosis, and non-cancer death.

##### Cancer Progression

The time to cancer progression from the moment the individual attains their current cancer stage is sampled as an exponential random variable with a rate given by the mean dwell time of the cancer type and current cancer stage. **Table S3** presents base case dwell times synthesized from published literature and expert surveys.^5,6^

**Table S3.** Cancer type- and stage-specific dwell times.

|  | **Mean dwell time (years)** | | | |
| --- | --- | --- | --- | --- |
| **Cancer type** | **Stage I** | **Stage II** | **Stage III** | **Stage IV** |
| **Breast** | 3 | 2 | 1 | 0.5 |
| **Cervical** | 4 | 2.5 | 1 | 0.75 |
| **Colorectal** | 1 | 1.5 | 1.25 | 0.75 |
| **Endometrial** | 3.5 | 2.25 | 1 | 0.5 |
| **Esophageal** | 2 | 1.5 | 1 | 1 |
| **Gastric** | 0.75 | 1 | 1 | 0.5 |
| **Head and Neck** | 2.5 | 1.5 | 1.25 | 0.5 |
| **Kidney** | 4 | 2 | 1 | 0.5 |
| **Liver** | 2 | 1 | 0.5 | 0.5 |
| **Lung** | 2 | 1.5 | 1 | 1 |
| **Ovary** | 2 | 1.25 | 0.75 | 0.5 |
| **Pancreatic** | 1 | 1 | 0.75 | 0.5 |
| **Prostate** | 7 | 5 | 3 | 1.5 |
| **Urinary Bladder** | 5.5 | 5.5 | 4.5 | 1 |

##### SoC Diagnosis

The time to SoC diagnosis from the moment the individual attains their current cancer stage is sampled as an exponential random variable with a rate given by the incidence rate for their (sex, race, age group, cancer type, cancer stage). Diagnosis was assumed to occur immediately upon advancement to stage IV cancer due to the high likelihood of having symptoms requiring medical care.

##### MCED Diagnosis

MCED testing is administered at deterministic time points as determined by the testing interval (*I*). The prevalent round of MCED testing occurs at model time $T=0$. For $T>0$ and not integer, representing the time at which an individual attains a new cancer stage, the next time point at which testing occurs is given by $T^{'}=\left\lceil T\div I \right\rceil\times I$, where $\left\lceil\cdot\right\rceil$ is the ceiling operator, following by testing at $T^{'}+I$, $T^{'}+2I$, $T^{'}+3I$, etc. The individual must be aged <85 years at the time of testing. The probability of the MCED test detecting cancer is given by the sensitivity for the cancer type and current cancer stage. Test sensitivities were derived from a large, multi-center, prospective, case-control study.^7^

##### Cancer Death

The time of cancer death (or non-cancer death) after diagnosis is determined by following survival curves. The full methodology is detailed in **Section S5**.

### S4 Calibration

Population-level cancer registries report observed cancer cases but do not characterize the volume of undiagnosed disease. Therefore, the prevalence and total incidence of cancer may be much higher than what is observed in registries. A backwards induction approach was developed to estimate the unobserved cancer burden.^8,9^ Using the rationale that cancer is a progressive disease where a case of late-stage cancer must have existed at an earlier time point as a case of early-stage cancer, stage IV cases were backtracked to stages I-III based on dwell times. From this, we estimated the unobserved cancer prevalence and incidence for each combination of cancer type, cancer stage, sex, race, and age.

For each cancer type, we used the outputs from the unobserved incidence methodology as initial estimates for the rate of oncogenesis, the initial prevalence by stage, and the rate of SoC diagnosis by stage. These parameters were subsequently calibrated at the cancer type and stage level, maintaining the original ratios between different combinations of sex, race, and age. The calibration target was annual incidence rates of diagnosis averaged over calendar years 2015–2021.^2^

Calibration was performed on an open cohort version of the model where individuals aged 40–49 years were also initialized and “entered” the model when they attained 50 years of age. For example, an individual initialized at age 45 would enter the model at *T* = 5. Thus, the model replicates population dynamics that may influence cancer diagnosis rates over the calibration period.

**Figure S2** compares final model outputs against SEER-reported incidence.

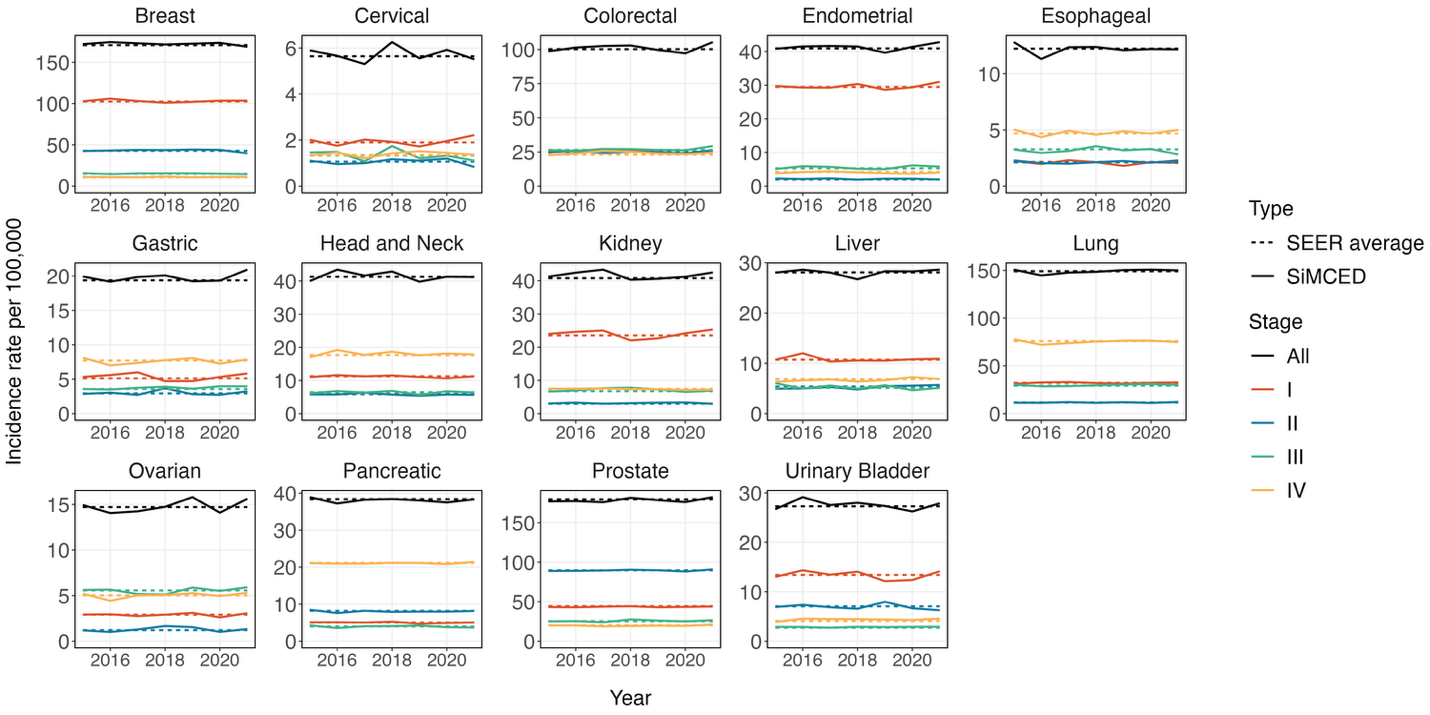

**Figure S2.** SEER-reported versus SiMCED-generated incidence by cancer type and stage. Abbreviations: SEER, Surveillance, Epidemiology, and End Results; SiMCED, Simulation Model for MCED.

### S5 Cancer Mortality

Before diagnosis, an individual with cancer cannot die of cancer. After diagnosis, the patient follows survival curves to determine the time of death. However, this means earlier diagnosis due to MCED testing can lead to earlier death. To avoid artificially penalizing earlier diagnosis, we enforce the following logic to delay the risk of death until the time of death in “SoC”:

- If the cause of death in “SoC” was cancer, then the patient is kept alive until the time of cancer death in “SoC”. After that, they follow the survival curve of the stage at diagnosis in “SoC + MCED” starting in year

$\lfloor($time of cancer death in “SoC” $)-$ $($time of MCED diagnosis in “SoC + MCED”$)\rfloor$,

where $\lfloor\cdot\rfloor$ is the floor operator. In other words, it is assumed that the patient survived the initial portion of the survival curve up to the time of cancer death in “SoC”.

- If the counterfactual cause of death in “SoC” was non-cancer, then the cause of death in “SoC + MCED” will also be non-cancer at the same time as “SoC”. Thus, earlier detection of cancer does not affect non-cancer death.

**Figure S3A** summarizes the above logic as a decision tree; **Figure S3B** presents an example application of this logic.

Cancer survival is evaluated each year after diagnosis on a discrete time basis (see the end of **Section S6**). This is to give the model the flexibility to use empirical non-parametric survival curves. If death occurs in a given year, a random perturbation is applied to uniformly distribute the time of death across the year interval. This means that “SoC + MCED” death can occur earlier in the same year as “SoC” death (due to the sampling of a smaller random perturbation). In this situation, the time of death is set to be identical to that of “SoC” death, without changing the type of death.

**(A)**

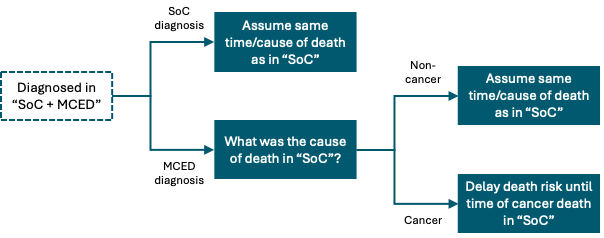

**(B)**

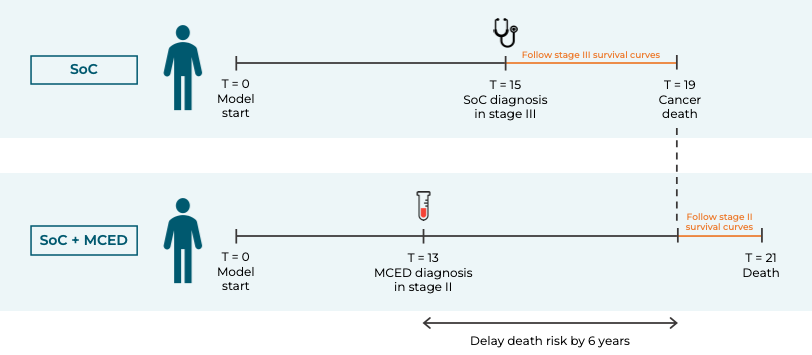

**Figure S3.** **(A)** Post diagnosis mortality decision tree. **(B)** Example application. Abbreviations: MCED, multi-cancer early detection, SoC, standard of care.

### S6 Survival Analysis

##### Data Source

The input data was summary survival curves for calendar years 2010–2021 stratified by cancer type, cancer stage, and 5-year age group from the SEER database.^2^ To simultaneously model cancer and non-cancer mortality, both observed and relative survival were considered. Observed survival describes the time to any-cause death; relative survival describes the time to cancer death.

##### Construction of Patient-Level Data

We loosely followed the methodology described in Hoyle et al. (2011) to construct patient-level data assuming no censorship except at the end of the observation period.^10^ We performed survival model fitting on the patient-level data using R package flexsurv. We considered the following parametric models: exponential (exp), Gompertz (gompertz), log-logistic (llogis), Weibull ­(accelerated failure time) (weibull) and Weibull (proportional hazards) (weibullPH). Separate models were fitted for each combination of cancer type, cancer stage, and age group (but the same model for observed and relative survival), and extrapolated up to 50 years. We followed NICE DSU guidelines for survival model evaluation,^11^ as outlined below.

##### Log-Cumulative Hazard Plots

As an initial validation step, we constructed log-cumulative hazard plots, i.e., $-\log\left( \log S\left( t \right) \right)$ versus $\log t$, where $S\left( t \right)$ denotes the SEER survival curve, to examine the empirical hazard rate over time. Since hazard rates were generally not time-constant, the use of the exponential survival model was ruled out.

##### AIC and BIC Test

Model selection based on AIC and BIC was inconclusive because differences were small. Moreover, visual inspection determined that the “non-optimal” models also produced plausible extrapolations. Internal validation was needed to identify the model with the best predictive power.

##### Internal Validation

We refitted survival models to calendar years 2010–2018 of the SEER survival curves and compared projections for 2019–2021 to the actual data points for those years. The model that minimized the sum of absolute errors (combining observed and relative survival) was chosen as the final model.

##### Model Selection Logic

In order to avoid situations where the curvature abruptly changes between consecutive age groups, we enforced several logical rules in the following order:

1. If age group $A$ uses model $X$ but age groups $\left( A -1 \right)$ and $\left( A+1 \right)$ both use model $Y$, then age group $A$ will switched to model $Y$, for $A\in\{$55–59, 60–64, 65–69, 70–74, 75–59, 80–84$\}$.
2. Age group 50–54 should use the same model as age group 55–59.
3. Age group 85+ should use the same model as age group 80–84.

##### Conversion to Annual Probabilities of Death

Let $S_{\mathrm{obs}}\left( t \right)$ and $S_{\mathrm{rel}}\left( t \right)$ denote the optimal fitted and extrapolated observed and relative survival curves, respectively, with $S_{\mathrm{obs}}\left( 0 \right)=S_{\mathrm{rel}}\left( 0 \right)=1$. Then,

$$p_{\mathrm{obs}}\left( t \right)=\frac{S_{\mathrm{obs}}\left( t-1 \right)-S_{\mathrm{obs}}\left( t \right)}{S_{\mathrm{obs}}\left( t-1 \right)}$$

and

$$p_{\mathrm{rel}}\left( t \right)=\frac{S_{\mathrm{rel}}\left( t-1 \right)-S_{\mathrm{rel}}\left( t \right)}{S_{\mathrm{rel}}\left( t-1 \right)} \cdot\prod_{s=1}^{t-1} \frac{1}{1-\left( p_{\mathrm{obs}}\left( s \right)-p_{\mathrm{rel}}\left( s \right) \right)} .$$

Note that $p_{\mathrm{rel}}\left( t \right)$ needs to be inflated by the product term to account for the loss of patients due to non-cancer mortality over the previous years.

##### Model Implementation

In year $t$ after diagnosis, a random uniform variate $U$ is generated. If $U<p_{\mathrm{rel}}\left( t \right)$, then the patient experiences cancer death in year $t$. If $p_{\mathrm{rel}}\left( t \right)<U<p_{\mathrm{obs}}\left( t \right)$, then the patient experiences non-cancer death in year $t.$ Otherwise, the patient survives year $t$. This process is repeated in each year $t$ until death occurs.

**SUPPLEMENTARY RESULTS**

***Table S4.*** *Distribution of LYG by cancer type and downstaging flow.*

| Stratification | LYG = 0 | LYG > 0 | LYG > 1 | LYG > 5 | LYG > 10 |
| --- | --- | --- | --- | --- | --- |
| By cancer type |  |  |  |  |  |
| Breast | 95% | 5% | 5% | 3% | 0% |
| Cervical | 61% | 39% | 35% | 25% | 0% |
| Colorectal | 78% | 22% | 20% | 13% | 0% |
| Endometrial | 93% | 7% | 6% | 4% | 0% |
| Esophageal | 72% | 28% | 21% | 11% | 0% |
| Gastric | 67% | 33% | 27% | 16% | 0% |
| Head and Neck | 78% | 22% | 19% | 12% | 0% |
| Kidney | 89% | 11% | 10% | 7% | 0% |
| Liver | 66% | 34% | 26% | 13% | 0% |
| Lung | 72% | 28% | 21% | 10% | 0% |
| Ovarian | 84% | 16% | 12% | 5% | 0% |
| Pancreatic | 58% | 42% | 29% | 11% | 0% |
| Prostate | 100% | 0% | 0% | 0% | 0% |
| Urinary Bladder | 77% | 23% | 19% | 12% | 0% |
| By downstaging flow |  |  |  |  |  |
| I to I | 99% | 1% | 1% | 1% | 0% |
| II to I | 59% | 41% | 34% | 22% | 0% |
| II to II | 96% | 4% | 3% | 2% | 0% |
| III to I | 39% | 61% | 53% | 37% | 0% |
| III to II | 50% | 50% | 41% | 23% | 0% |
| III to III | 83% | 17% | 13% | 5% | 0% |
| IV to I | 18% | 82% | 74% | 56% | 0% |
| IV to II | 16% | 84% | 72% | 45% | 0% |
| IV to III | 22% | 78% | 61% | 30% | 0% |
| IV to IV | 100% | 0% | 0% | 0% | 0% |
| All | 84% | 16% | 13% | 7% | 0% |

### *Abbreviations: LYG, life-years gained.*

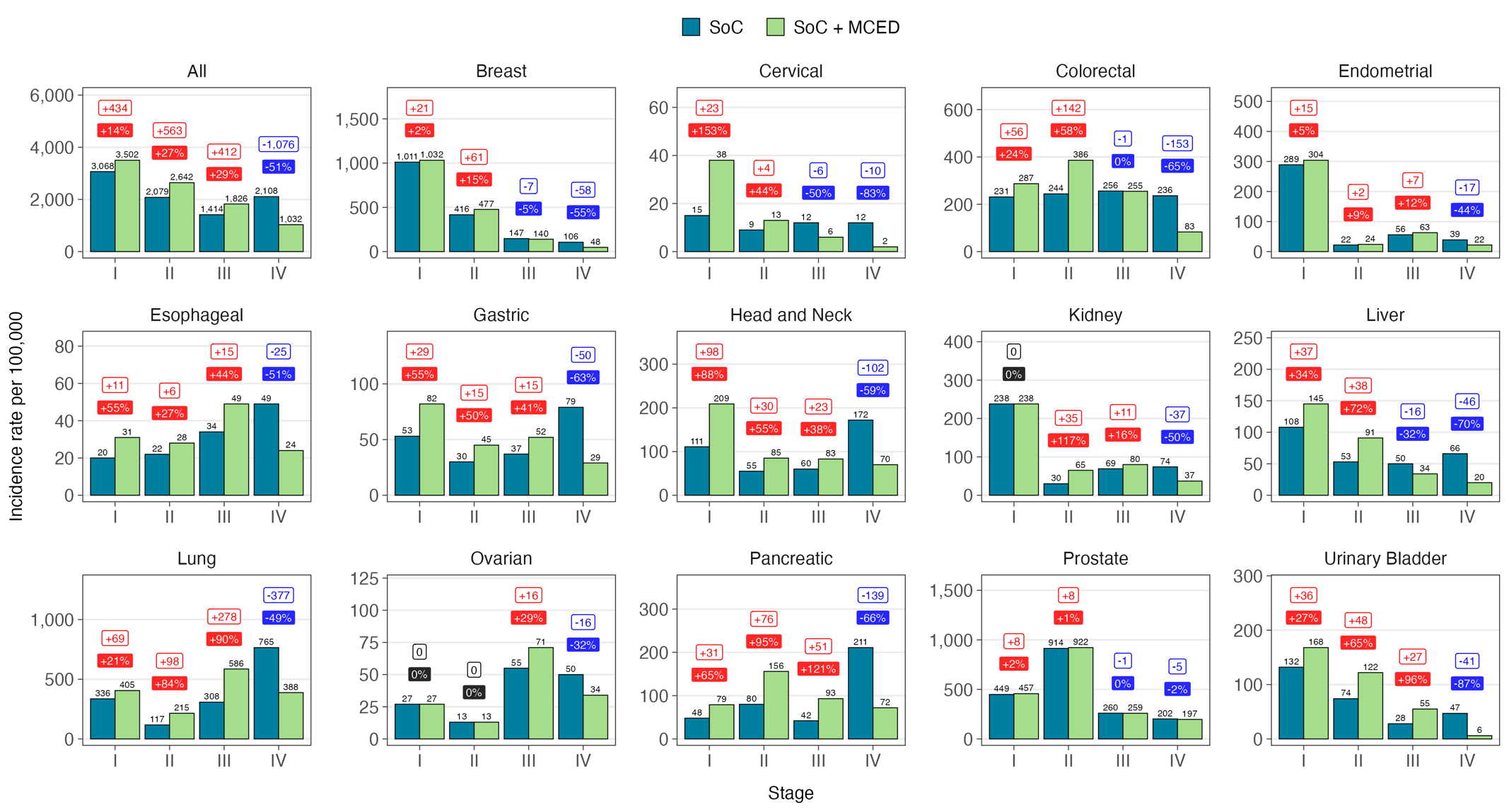

***Figure S4.*** *10-year stage shift by cancer type. Abbreviations: MCED, multi-cancer early detection; SoC, standard of care.*

**REFERENCES**

1.	Chhatwal J, Xiao J, ElHabr AK, et al. The impact of multicancer early detection tests on cancer stage shift: A 10-year microsimulation model. *Cancer*. 2025;131(22):e70075. doi:10.1002/cncr.70075

2.	National Cancer Institute, DCCPS, Surveillance Research Program. Surveillance Research Program, National Cancer Institute SEER*Stat software. Published online November 2023. seer.cancer.gov/seerstat

3.	Centers for Disease Control and Prevention (CDC). Single-Race Population Estimates. https://wonder.cdc.gov/single-race-population.html

4.	Arias E, Xu J, Tejada-Vera B, Bastian B. U.S. State Life Tables, 2021. *National Vital Statistics Reports*. 2024;73(7).

5.	Broder MS, Ailawadhi S, Beltran H, et al. Estimates of stage-specific preclinical sojourn time across 21 cancer types. *JCO*. 2021;39(15_suppl):e18584-e18584. doi:10.1200/JCO.2021.39.15_suppl.e18584

6.	Shah N, Hathaway C, Tyson C, Cohain A, Li Y. MSR64 Novel Empirical Methods to Derive Stage-Specific Dwell Time and Implications for Multi-Cancer Early Detection (MCED) Modeling. *Value in Health*. 2022;25(7):S530. doi:10.1016/j.jval.2022.04.1271

7.	Gainullin V, Bae J, Guthrie VB, et al. Abstract A056: Performance of multi-biomarker class reflex testing in a prospectively-collected cohort. *Clinical Cancer Research*. 2024;30(21_Supplement):A056. doi:10.1158/1557-3265.LIQBIOP24-A056

8.	Chhatwal J, ElHabr A, Tyson C, et al. Correlation of unobserved incidence of cancer in earlier stages with the observed incidence. *Journal of Clinical Oncology*. 2023;41(16_suppl):10634-10634. doi:10.1200/JCO.2023.41.16_suppl.10634

9.	ElHabr A, Tyson C, Cao X, et al. EPH232 The Large Hidden Prevalence Rate of Cancer Using Backward Induction Method Reveals Screening Opportunity in Earlier Stages. *Value in Health*. 2023;26:S205. doi:10.1016/j.jval.2023.03.2578

10.	Hoyle MW, Henley W. Improved curve fits to summary survival data: application to economic evaluation of health technologies. *BMC Medical Research Methodology*. 2011;11(1):139. doi:10.1186/1471-2288-11-139

11.	Latimer N. *NICE DSU TECHNICAL SUPPORT DOCUMENT 14: SURVIVAL ANALYSIS FOR ECONOMIC EVALUATIONS ALONGSIDE CLINICAL TRIALS - EXTRAPOLATION WITH PATIENT-LEVEL DATA*.; 2013. https://sheffield.ac.uk/nice-dsu/tsds/survival-analysis
